## Supplementary data for "Identifying healthy individuals with Alzheimer neuroimaging phenotypes in the UK Biobank"

[1]Department of Computer Science and Technology, University of Cambridge, UK
[2]Brain Mapping Unit, Department of Psychiatry, University of Cambridge, UK
[3]Autism Research Centre, Department of Psychiatry, University of Cambridge, UK
[4]Department of Clinical Neurosciences and Cambridge University Hospitals NHS Trust, University of Cambridge, UK
[*]
[**]Data used in preparation of this article were obtained from the Alzheimer's Disease Neuroimaging Initiative (ADNI) database (adni.loni.usc.edu). As such, the investigators within the ADNI contributed to the design and implementation of ADNI and/or provided data but did not participate in analysis or writing of this report. A complete listing of ADNI investigators can be found at: http://adni.loni.usc.edu/wp-content/uploads/how_to_apply/ADNI_Acknowledgement_List.pdf

### Comparison between monte carlo dropout and single forward pass

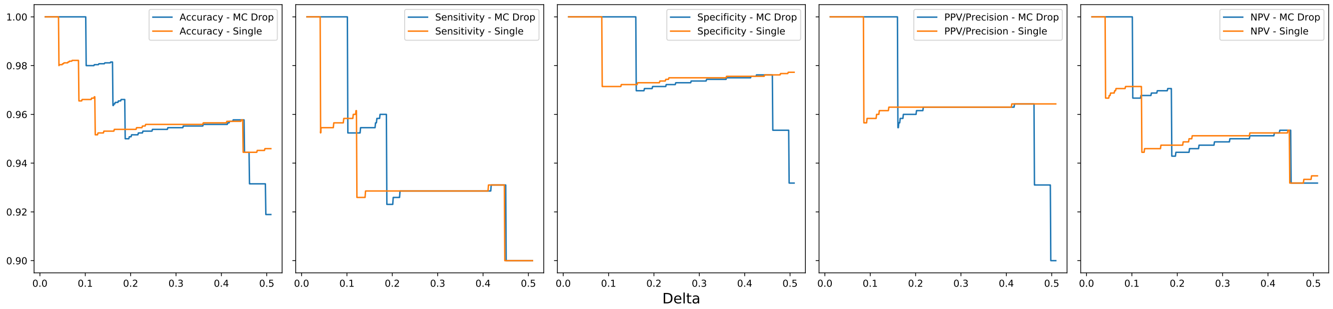


**Figure S1. Comparison between our model with Monte Carlo dropout (with 50 samples), and a corresponding single forward pass one, for the ADNI test set.** We sequentially evaluate performance by including different number people according to the model's output, where delta corresponds to the distance to the model's extremes (ie. 0 or 1). As delta increases, more people are included with an output closer to 0.5. Metrics are calculated for a cut-off of 0.5.

### NACC clinical scores

### Validation of the an AD cut-off score of 0.5

We investigated the association of AD score with clinical scores using piecewise linar regression models both with an without a breakpoint, and a linear regression model with no breakpoint. In the flexible breakpoint model, the breakpoint was restricted to between 0.25 and 0.75 to avoid improbable extreme values. We report the comparisons in model fit between the models. For each comprison one of the models 'wins', indicated in bold in table S7 with a model fit (Expected Log Probability Density Function, ELPDF) score of 0. The difference in model fit is expressed as a negative ELPDF. A model can be considered substantially worse if the magnitude of the difference is large and the standard deviation of the ELPDF is substantially smaller than the difference in model fit. This pattern is seen in MMSE, MoCA and semantic fluency for the linear regression model without a breakpoint, demonstrating that the two piecewise regression models are broadly equivalent, but both piecewise regression models are a substantially better fit than the simple linear regression model without a breakpoint.

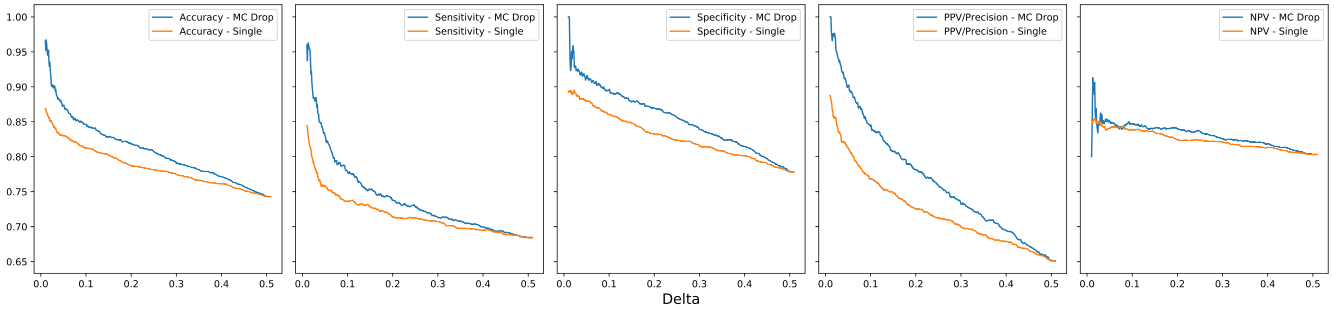


**Figure S2. Comparison between our model with Monte Carlo dropout (with 50 samples), and a corresponding single forward pass one, for the NACC dataset with only AD and controls.** We sequentially evaluate performance by including different number people according to the model's output, where delta corresponds to the distance to the model's extremes (ie. 0 or 1). As delta increases, more people are included with an output closer to 0.5. Metrics are calculated for a cut-off of 0.5. Our model consistently outperforms the corresponding neural network with one forward pass.

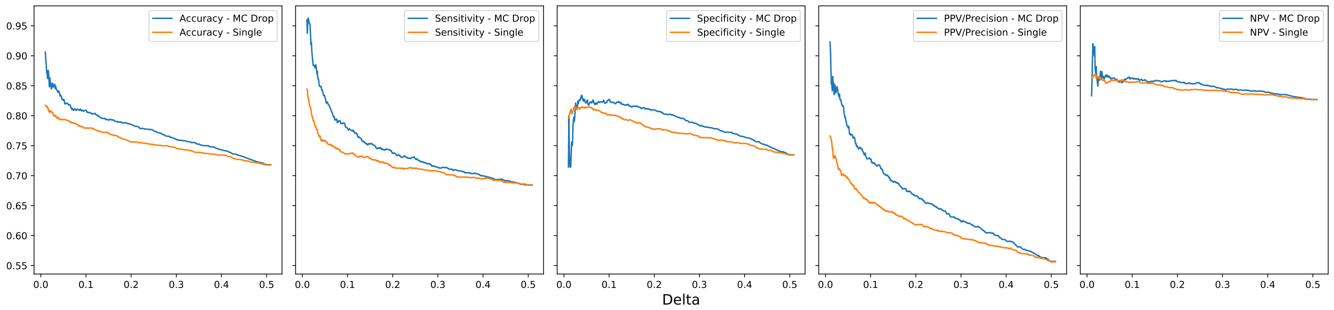


**Figure S3. Comparison between our model with Monte Carlo dropout (with 50 samples), and a corresponding single forward pass one, for the NACC dataset including all diagnoses.** We sequentially evaluate performance by including different number people according to the model's output, where delta corresponds to the distance to the model's extremes (ie. 0 or 1). As delta increases, more people are included with an output closer to 0.5. Metrics are calculated for a cut-off of 0.5. Our model consistently outperforms the corresponding neural network with one forward pass.

There was a slightly better model fit in all cases for a variable rather than fixed breakpoint, however there was only weak evidence for this given that the standard deviation for the expected log probability density function was approximately similar to the difference in model fit. Reassuringly, there was no convincing superiority of the piecewise regression models over linear regression models in forward and backward digit span (that did not show differences between AD positive and negative scores), suggesting that the piecewise regression models did not overfit the data.

**Table S1.** The association between NACC MMSE scores and AD scores with a variable breakpoint.

| | mean | se_mean | sd | 2.5% | 25% | 50% | 75% | 97.5% | n_eff | Rhat |
|---|---|---|---|---|---|---|---|---|---|---|
| intercept | 0.081 | 0.0027 | 0.097 | -0.11 | 0.016 | 0.08 | 0.15 | 0.27 | 1332 | 1.00 |
| bp | 0.67 | 0.0021 | 0.084 | 0.43 | 0.64 | 0.7 | 0.73 | 0.75 | 1689 | 1.00 |
| slope_before | -0.19 | 0.0017 | 0.069 | -0.33 | -0.24 | -0.19 | -0.15 | -0.054 | 1628 | 1.00 |
| slope_after | -1.4 | 0.0053 | 0.22 | -1.8 | -1.5 | -1.3 | -1.2 | -0.94 | 1705 | 1.00 |
| slope_difference | -1.2 | 0.0066 | 0.26 | -1.7 | -1.3 | -1.2 | -0.98 | -0.65 | 1585 | 1.00 |
| error | 0.87 | 0.00058 | 0.028 | 0.82 | 0.85 | 0.87 | 0.88 | 0.92 | 2247 | 1.00 |

**Table S2.** The association between NACC MMSE scores and AD score with a breakpoint of the AD score fixed at 0.5

| | mean | se_mean | sd | 2.5% | 25% | 50% | 75% | 97.5% | n_eff | Rhat |
|---|---|---|---|---|---|---|---|---|---|---|
| intercept | 0.15 | 0.0026 | 0.096 | -0.036 | 0.083 | 0.14 | 0.21 | 0.34 | 1316 | 1.00 |
| slope_before | -0.17 | 0.0021 | 0.077 | -0.32 | -0.23 | -0.17 | -0.12 | -0.022 | 1350 | 1.00 |
| slope_after | -1.1 | 0.004 | 0.15 | -1.4 | -1.2 | -1.1 | -1 | -0.82 | 1418 | 1.00 |
| slope_difference | -0.95 | 0.006 | 0.21 | -1.4 | -1.1 | -0.94 | -0.81 | -0.53 | 1274 | 1.00 |
| error | 0.87 | 0.00059 | 0.026 | 0.82 | 0.85 | 0.87 | 0.89 | 0.92 | 1970 | 1.00 |

**Table S3.** A linear model of NACC MMSE scores and AD scores with no breakpoint.

| | mean | se_mean | sd | 2.5% | 25% | 50% | 75% | 97.5% | n_eff | Rhat |
|---|---|---|---|---|---|---|---|---|---|---|
| intercept | 0.081 | 0.0027 | 0.097 | -0.11 | 0.016 | 0.08 | 0.15 | 0.27 | 1332 | 1.00 |
| bp | 0.67 | 0.0021 | 0.084 | 0.43 | 0.64 | 0.7 | 0.73 | 0.75 | 1689 | 1.00 |
| slope_before | -0.19 | 0.0017 | 0.069 | -0.33 | -0.24 | -0.19 | -0.15 | -0.054 | 1628 | 1.00 |
| slope_after | -1.4 | 0.0053 | 0.22 | -1.8 | -1.5 | -1.3 | -1.2 | -0.94 | 1705 | 1.00 |
| slope_difference | -1.2 | 0.0066 | 0.26 | -1.7 | -1.3 | -1.2 | -0.98 | -0.65 | 1585 | 1.00 |
| error | 0.87 | 0.00058 | 0.028 | 0.82 | 0.85 | 0.87 | 0.88 | 0.92 | 2247 | 1.00 |

**Table S4.** Caption

| | **Model difference** | **SE difference** |
|---|---|---|
| Fixed breakpoint | 0.0 | 0.0 |
| Variable breakpoint | -1.3 | 1.0 |
| No breakpoint | -10.0 | 5.3 |

**Table S5.** The association between NACC MoCA scores and AD scores with a variable breakpoint.

| | mean | se_mean | sd | 2.5% | 25% | 50% | 75% | 97.5% | n_eff | Rhat |
|---|---|---|---|---|---|---|---|---|---|---|
| intercept | 0.081 | 0.0027 | 0.097 | -0.11 | 0.016 | 0.08 | 0.15 | 0.27 | 1332 | 1.00 |
| bp | 0.67 | 0.0021 | 0.084 | 0.43 | 0.64 | 0.7 | 0.73 | 0.75 | 1689 | 1.00 |
| slope_before | -0.19 | 0.0017 | 0.069 | -0.33 | -0.24 | -0.19 | -0.15 | -0.054 | 1628 | 1.00 |
| slope_after | -1.4 | 0.0053 | 0.22 | -1.8 | -1.5 | -1.3 | -1.2 | -0.94 | 1705 | 1.00 |
| slope_difference | -1.2 | 0.0066 | 0.26 | -1.7 | -1.3 | -1.2 | -0.98 | -0.65 | 1585 | 1.00 |
| error | 0.87 | 0.00058 | 0.028 | 0.82 | 0.85 | 0.87 | 0.88 | 0.92 | 2247 | 1.00 |

**Table S6.** The association between NACC MMSE scores and AD score with a breakpoint of the AD score fixed at 0.5

| | mean | se_mean | sd | 2.5% | 25% | 50% | 75% | 97.5% | n_eff | Rhat |
|---|---|---|---|---|---|---|---|---|---|---|
| intercept | 0.15 | 0.0026 | 0.096 | -0.036 | 0.083 | 0.14 | 0.21 | 0.34 | 1316 | 1.00 |
| slope_before | -0.17 | 0.0021 | 0.077 | -0.32 | -0.23 | -0.17 | -0.12 | -0.022 | 1350 | 1.00 |
| slope_after | -1.1 | 0.004 | 0.15 | -1.4 | -1.2 | -1.1 | -1 | -0.82 | 1418 | 1.00 |
| slope_difference | -0.95 | 0.006 | 0.21 | -1.4 | -1.1 | -0.94 | -0.81 | -0.53 | 1274 | 1.00 |
| error | 0.87 | 0.00059 | 0.026 | 0.82 | 0.85 | 0.87 | 0.89 | 0.92 | 1970 | 1.00 |

**Table S7.** ELPDF = Expected Log Probability Density Function, sd = Standard deviation.

| | **Fixed breakpoint (0.5)** | **Variable breakpoint** | | **Linear model** |
|---|---|---|---|---|
| | **ELPDF (sd)** | **Breakpoint** | **ELPDF (sd)** | **ELPDF (sd)** |
| MMSE | -1.3 (1.0) | 0.67 | **0.0 (0)** | -10.0 (5.3) |
| MoCA | -0.5 (0.6) | 0.60 | **0.0 (0)** | -12.4 (6.8) |
| Digit span (forward) | -0.4 (0.3) | 0.58 | **0.0 (0)** | -2.6 (2.6) |
| Digit span (backward) | -0.2 (0.2) | 0.56 | **0.0 (0)** | -0.5 (1.6) |
| Semantic fluency | -0.5 (0.4) | 0.59 | **0.0 (0)** | -9.6 (4.5) |
| Trails B | **0.0 (0.2)** | 0.44 | **0.0 (0)** | -2.7 (2.8) |
| WAIS | **0.0 (0)** | 0.53 | -0.1 (0.1) | -0.3 (1.4) |
| Boston naming task | -1.7 (0.7) | 0.66 | **0.0 (0)** | -9.6 (4.6) |
